# Epigenome-wide association study for dilated cardiomyopathy in left ventricular heart tissue identifies putative gene sets associated with cardiac development and early indicators of cardiac risk

**DOI:** 10.1101/2024.07.16.24310537

**Authors:** Konstanze Tan, Darwin Tay, Wilson Tan, Hong Kiat Ng, Eleanor Wong, Michael P Morley, Gurpreet K Singhera, Pritesh R Jain, Fei Li Tai, Paul J Hanson, Thomas P Cappola, Kenneth B Margulies, Roger Foo, Marie Loh

## Abstract

**Background:** Methylation changes linked to dilated cardiomyopathy (DCM) affect cardiac gene expression. We investigate DCM mechanisms regulated by CpG methylation using multi-omics and causal analyses in the largest cohort of left ventricular tissues available.

**Methods:** We mapped DNA methylation at ∼850,000 CpG sites, performed array-based genotyping and RNA sequencing in left-ventricular tissue samples from failing and non-failing hearts across two independent DCM cohorts (discovery n=329, replication n=85). Summary data-based Mendelian Randomization (SMR) was applied to explore the causal contribution of sentinel CpGs to DCM. Fine-mapping of regions surrounding sentinel CpGs revealed additional signals for cardiovascular disease risk factors. Coordinated changes across multiple CpG sites were examined using weighted gene correlation network analysis (WGCNA).

**Results:** We identified 194 epigenome-wide significant CpGs associated with DCM (discovery P<5.96E-08), enriched in active chromatin states in heart tissue. Amongst these, 183 sentinel CpGs significantly influenced the expression of 849 proximal genes (±1Mb). SMR suggested the causal contribution of two sentinel CpGs to DCM and 36 sentinel CpGs to the expression of 43 unique proximal genes (P<0.05). Colocalization analyses indicated that a single causal variant may underlie the methylation-gene expression relationship for three sentinel CpGs. Fine-mapping revealed additional signals linked to cardiovascular traits including hsCRP and blood pressure. Co-methylation modules were enriched in genes related to cardiac physiological and pathological processes and their corresponding transcriptional regulators.

**Conclusions:** Using the largest series of left ventricular tissue to date, this study investigates the causal role of cardiac methylation changes in DCM and suggests targets for experimental studies to probe DCM pathogenesis.

## INTRODUCTION

Dilated cardiomyopathy (DCM) is the most common form of cardiomyopathy and a leading indication for heart transplantation, affecting 1 in 250-400 individuals, with an incidence of 5-7 cases per 100,000 persons per year.^1^ DCM is characterized by a thinning and weakening of the left ventricular heart walls, with resulting contractile dysfunction.^2–4^ Clinical diagnostic criteria for DCM include a left ventricular ejection fraction (LVEF) of <45% and a left ventricular end-diastolic dimension >112% of that predicted for age and body surface area.^5^ There is marked heterogeneity in both the prognosis and age of onset for DCM, with most patients becoming symptomatic between 20-50 years of age.^6^ DCM accounts for a substantial 2-3% of yearly sudden cardiac death (SCD) events in which the heart stops beating, resulting in a sudden loss of blood flow to vital organs.^7^

Over the past decade, the genetic basis for DCM has only been partially unraveled.^8^ Beyond genetic predisposition, environmental factors could influence the incidence and course of DCM.^4^ DNA methylation, a key regulator of gene expression, as well as a molecular integrator of genetic and environmental influences, has been investigated in DCM to gain insights into its molecular pathogenesis. To date, multiple epigenome-wide association studies (EWAS) of DCM have identified a handful of differentially methylated loci.^9–14^ This includes the reporting of conservation in methylation patterns between myocardium and peripheral blood at the *NPPA* and *NPPB* loci encoding cardiac stress markers, which are currently gold standard indicators for heart failure prognosis in clinics, thereby demonstrating the potential of DNA methylation as a novel class of DCM biomarkers. However, these epigenome-wide methylation studies of DCM have largely been limited by sample size (n=8-72) owing to the scarcity of left ventricular tissue.

In this study, we sought to expand our understanding of myocardial-specific methylation changes in DCM by analyzing intra-operative left ventricular (LV) tissue from a cohort of 329 individuals of White and African American ancestries (n = 159 cases, 170 controls). This study represents the largest investigation of DCM methylation conducted in myocardial tissue to date. Through integrative omics, causal analyses and fine-mapping of putative methylation markers, we aim to elucidate the putative causal involvement of methylation alterations to DCM pathogenesis.

## METHODS

### Description of samples

For each cohort, written informed consent for the research use of donated left ventricular (LV) tissue were obtained. For heart transplant recipients, consent was obtained from the transplant recipient. For brain-dead organ donors, consent was obtained from the next-of-kin. All analyses and study protocols were approved by the relevant institutional review boards.

### Myocardial Applied Genomics Network Cohort (MAGNet

LV free-wall tissue was harvested at the time of cardiac surgery from subjects with heart failure undergoing transplantation and from donor hearts deemed unsuitable for transplantation but with apparently normal ventricular function. Hearts were perfused immediately with cold in-situ high-potassium cardioplegia before cardiectomy to arrest contraction and prevent ischemic damage. Tissue samples were promptly frozen in liquid nitrogen. Only samples diagnosed with ‘idiopathic DCM’ were analysed.

### Bruce McManus Cardiovascular Biobank Cohort (BMCB)

LV free-wall tissue (apical portion) was obtained from biopsies of patients with DCM. Biopsies of 1- to 2-mm diameter were washed in ice-cold saline (0.9% NaCl) and stored in liquid nitrogen until DNA extraction. Control tissue samples were non-postmortem donor hearts recovered within 0-2 hours of circulatory death from the International Institute for the Advancement of Medicine (IIAM). Samples with confirmed or suspected non-idiopathic etiology for DCM (e.g., cardiotoxicity, inflammation, familial, myocarditis, ischemia) were excluded.

### The iHealth-T2D study

iHealth-T2D is a prospective study of Indian Asian men and women living in West London, and recruited in 2016. At enrolment all participants completed a structured assessment of cardiovascular and metabolic health, including (i) history for CVD such as myocardial infarction, angina, and coronary heart disease; (ii) indicators of CVD risk including blood pressure (systolic and diastolic), Framingham Coronary Heart Disease score, and hypertension/history of hypertension, and (iii) collection of blood samples for measurement of lipid profile and markers such as high-sensitivity C-reactive protein (hsCRP) and creatinine. Methylation profiling was performed using genomic DNA from peripheral blood collected at enrolment. The study is approved by the National Research Ethics Service and all participants gave written informed consent.

### Quantification of DNA methylation

Genomic DNA was bisulfite converted using the EZ DNA methylation kit according to the manufacturer’s instructions (Zymo Research). Case and control samples were randomized in all experiments. Bisulfite-converted genomic DNA was quantified using the Infinium MethylationEPIC BeadChip (EPIC array). Retrieval of bead intensity and Illumina background correction was performed using the *minfi* R package (version 1.40.0). Markers were excluded from analyses if they: (i) were non-CpG; (ii) were present on sex chromosomes; or (iii) had low call-rate (<95%). Samples were excluded from analyses if they: (i) had low call-rate (<98%); (ii) had mislabelled gender; or (iii) were present in duplicate. After filtering, the discovery cohort (MAGNet) had 838,624 autosomal CpG probes and 329 samples, while the replication cohort (BMCB) had 841,904 autosomal CpG probes and 85 samples available for analysis.

### Statistical analyses of epigenome-wide data

Epigenome-wide methylation data was analysed using the CPACOR pipeline (incorporating Control Probe Adjustment and reduction of global CORrelation) as previously described.^16^ Quantile-normalised marker intensity values were then used to calculate per-CpG methylation beta values. Principal component analysis (PCA) was performed on built-in control probe intensities to identify PCs accounting for ∼ 95% of control probe variation which were included as predictors in regression models to adjust for technical biases (MAGNet: first 10 PCs; BMCB: first 3 PCs). The association of each autosomal CpG site with DCM was tested using logistic regression, adjusting for age, gender and control probe PCs (Model: DCM ∼ Beta(QN) + age + gender + PCs_control-probes_). Within the MAGNet cohort, regression was performed separately for each ancestry group, followed by a trans-ancestry inverse variance-weighted meta-analysis using METAL. Test-statistic bias and inflation in association results was adjusted pre- and post-meta-analysis, using a Bayesian algorithm implemented in the *bacon* R package (version 1.30.0).^27^

### Enrichment analysis of genomic regulatory features

Sentinel CpGs were evaluated for enrichment in genomic regulatory features compared to 1000 permutations of EPIC array CpGs. Enrichment was determined through a one-tailed permutation test, computing a P value by comparing overlap counts of the test set (sentinel CpGs) with the distribution from the background. Enrichment in eQTMs and transcription factor binding sites (TFBS) was examined against EPIC array CpG sites matched by methylation levels and variability using a sliding-window approach, wherein adjustments were made to mean and standard deviation values until 1000 permuted sites were reached.^28^ Known TFBS (N=1,210) were obtained from the Remap 2022 database. Enriched TFs were classified as ‘Cardiac TF’ if expressed in humans and validated *in vivo*^29,30^, ‘Computational-based’ if predicted as cardiac TFs computationally due to disruption of TFBS near heart-failure linked loci^31,32^, or ‘non-cardiac TF’ if there is no suggestion for a cardiac role in humans in existing literature. eFORGE V2 software assessed sentinel CpG enrichment in learned chromatin states, DNAse 1 hotspots, and regions marked by core histone marks across various tissues and cell subsets within the Roadmap Epigenomic Consortium database.^33^ For constructing the background set, eFORGE V2 software matched EPIC array CpG sites to sentinel CpGs based on gene and CpG island annotations.

### RNA sequencing

Gene expression (RNAseq) data was obtained from the same left ventricular samples analysed for methylation, where available. Prior to downstream analyses, genes were removed if they: (i) had low read counts (<10 counts in the minimum group size of samples) or (ii) were present on sex chromosomes. Gene expression levels were normalized using TMM (Trimmed Mean of M-values), recommended for quantitative trait loci analyses.^34^ More details on the sequencing, alignment, filtering and normalization of RNAseq data are available in Supplementary Methods.

### Expression quantitative trait methylation (eQTM) analysis

Expression quantitative trait methylation (eQTM) analysis was conducted to identify significant associations between sentinel CpG methylation and proximal gene expression (+/- 1Mb of the sentinel CpG based on gene TSS). Gene expression was modelled against CpG methylation with adjustment for age, gender, ethnicity, RNA Integrity number (RIN) and the top 5 PEER factors which represent latent sources of variability in the gene expression data, possibly including technical and unknown confounders.^35^ Association testing was performed using a linear model implemented within the *MatrixEQTL* R package (version 2.3).^15^

### Methylation quantitative trait loci analysis (meQTL)

DNA samples were obtained from the same left ventricular samples analysed for methylation, where available (MAGNet). Genotyping was performed using the Affymetrix Genome-Wide SNP Array 6.0, as previously described.^36^ SNPs were removed from the current analyses if they: (i) were multiallelic; (ii) were present in duplicate); (iii) had low imputation quality (R2<0.3) or (iv) were rare (ancestry-specific MAF<0.05). Methylation quantitative trait loci (meQTL) analysis was conducted to identify significant associations between SNPs and sentinel CpG methylation (+/-500kb). CpG methylation was regressed against SNP dosage values with adjustment for age, gender, methylation array control probe PCs capturing 95% of control probe variation. Association testing was performed using a linear model implemented within the *MatrixEQTL* R package (version 2.3).^15^ Regression was performed separately for each ancestry group, followed by a trans-ancestry inverse variance-weighted meta-analysis using METAL.

### Causal analyses (Mendelian Randomisation and Colocalisation)

Summary data-based Mendelian Randomization (SMR) was performed to investigate the putative causal contribution of sentinel CpGs to DCM. SMR uses summary-level association statistics from independent association studies to compute a causal estimate for the influence of an exposure (e.g. sentinel CpG methylation) on an outcome (e.g. DCM disease status, or gene expression).^24^ The significance of the causal estimate is determined using the Wald Test. Separate SMR analyses were conducted to examine causal relationships between sentinel CpG methylation and: i. DCM ii. proximal gene expression (+/-1Mb). For each sentinel CpG, the lead meQTL (P<0.05) that was also analysed for association with the respective outcome was chosen to be the instrumental variable (IV) for assessing causal relationships. Genetic associations for DCM and gene expression were obtained from the largest published GWAS of DCM (n= 355,381, UKBioBank) and left ventricular cis-eQTL study (n=386; GTEx v8 release; SNPs within +/- 1Mb of gene transcription start sites), respectively. We validated SMR-significant associations using one-sample MR and conducted colocalisation analyses to evaluate the posterior probabilities of a shared causal variant for the assessed traits. One-sample Mendelian Randomization (MR) was performed with individual-level genotype, methylation, and outcome data (MAGNet) using the 2-stage least squares regression method implemented in the *AER* R package (version: 1.2-12). Colocalization analysis was conducted using the *coloc.abf* function in the *coloc* R package (version 5.2.3), based on a +/-500kb region around each sentinel CpG as this was the window size used for identifying meQTL.^37^

### Weighted gene correlation network analysis (WGCNA)

We employed Weighted Gene Correlation Network Analysis (WGCNA) to construct co-methylation modules using DCM-associated CpGs from discovery-stage EWAS.^38^ These CpGs were (i) associated with DCM (FDR P<0.05); (ii) replicated with consistent association direction; and (iii) displayed variability (methylation beta SD>0.02). Using 32,198 CpGs we built a signed consensus co-methylation network using the *blockwiseconsensusModules* function in the *WGCNA* R package (version 1.72-5), utilizing settings recommended for our sample size (soft thresholding power = 12, merge cut height = 0.25 and minimum module size = 30). Methylation levels within consensus modules were summarized as module eigengenes (ME) and tested for correlation with DCM separately by ancestry (Model: DCM ∼ ME + Age + Gender + 10PCs). Inverse variance-weighted meta-analysis combined ancestry-specific results to determine module associations with DCM. Additionally, a hypergeometric test was applied to assess module enrichment in DCM sentinel CpGs (P<0.05) against all CpGs used for network construction. Genes mapped to CpGs within co-methylation modules for overrepresentation of Gene Ontology (GO) terms and biological processes from KEGG and REACTOME databases using the *clusterProfiler* R package (version 4.10.0). GO gene sets were obtained from *org.Hs.eg.db* (version 3.18.0), while KEGG and REACTOME datasets were accessed via the *msigdbr* R package (version 7.5.1).

### Targeted methylation sequencing

Targeted methylation sequencing was performed using genomic DNA from peripheral blood samples collected from participants of the iHealth-T2D study. Targeted methylation sequencing was performed for regions defined as +/-500bp of DCM sentinel CpGs, supported by previous publications demonstrating a decay in correlation between methylation sites beyond 1-2kb,^39,40^ and from our in-house whole-genome bisulfite sequencing dataset where we observed that |r|>0.2 was within +/-500bp for most of the assessed CpGs (data unpublished). Additional details on probe design, sample processing and library preparation, sequencing and quality control steps employed are available in Supplementary Methods.

### Statistical analyses of targeted methylation sequencing data

Within each sequenced region, the association of each CpG site with CVD-related traits was tested using linear regression for continuous traits and logistic regression for binary traits, adjusting for age, gender and white blood cell proportion of six white blood cell sub-populations (CD8 T cells, CD4 T cells, Natural Killer Cells, B-cells, Monocytes, Granulocytes) estimated by the Houseman algorithm.^41^ In parallel, pairwise correlation in methylation levels between CpGs within regions was calculated using the R function *cor*, which computes the correlation coefficient. For a given pair of CpGs, only samples where methylation data was available for both CpGs being compared were used to calculate correlation.

### Construction of methylation score

We constructed a methylation score from CpGs in fine-mapped regions and examined the association of these scores with CVD traits. Effect sizes of association between individual CpG loci and CVD traits were used as weights for constructing the methylation score. Specifically, we defined trait-specific methylation scores within each region as a linear combination of *k* CpG site beta values *b* and weights *w*:

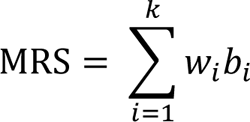

Within each region, the association of each MRS with CVD-related traits was assessed, adjusting for the same covariates as in single loci association testing described in **‘*Statistical analyses of targeted methylation sequencing data’*.**

## RESULTS

### Overview of study design

Our study design is summarised in Figure 1. In brief, we first carried out an epigenome-wide association investigation of DCM using 414 left ventricular samples obtained from two repositories: the Myocardial Applied Genomics network (MAGNet; n=329 [discovery]) and Bruce McManus Cardiovascular Biobank (BMCB; n=85 [replication]) (Supplementary Table 1). Integrative omics analyses were performed on the identified sentinel CpGs. To discover additional signals beyond CpG sites captured by the methylation array, we conducted fine-mapping of selected top-ranking loci in blood samples obtained from a population-based cohort (iHealth-T2D, n=1974).

**Figure 1.**
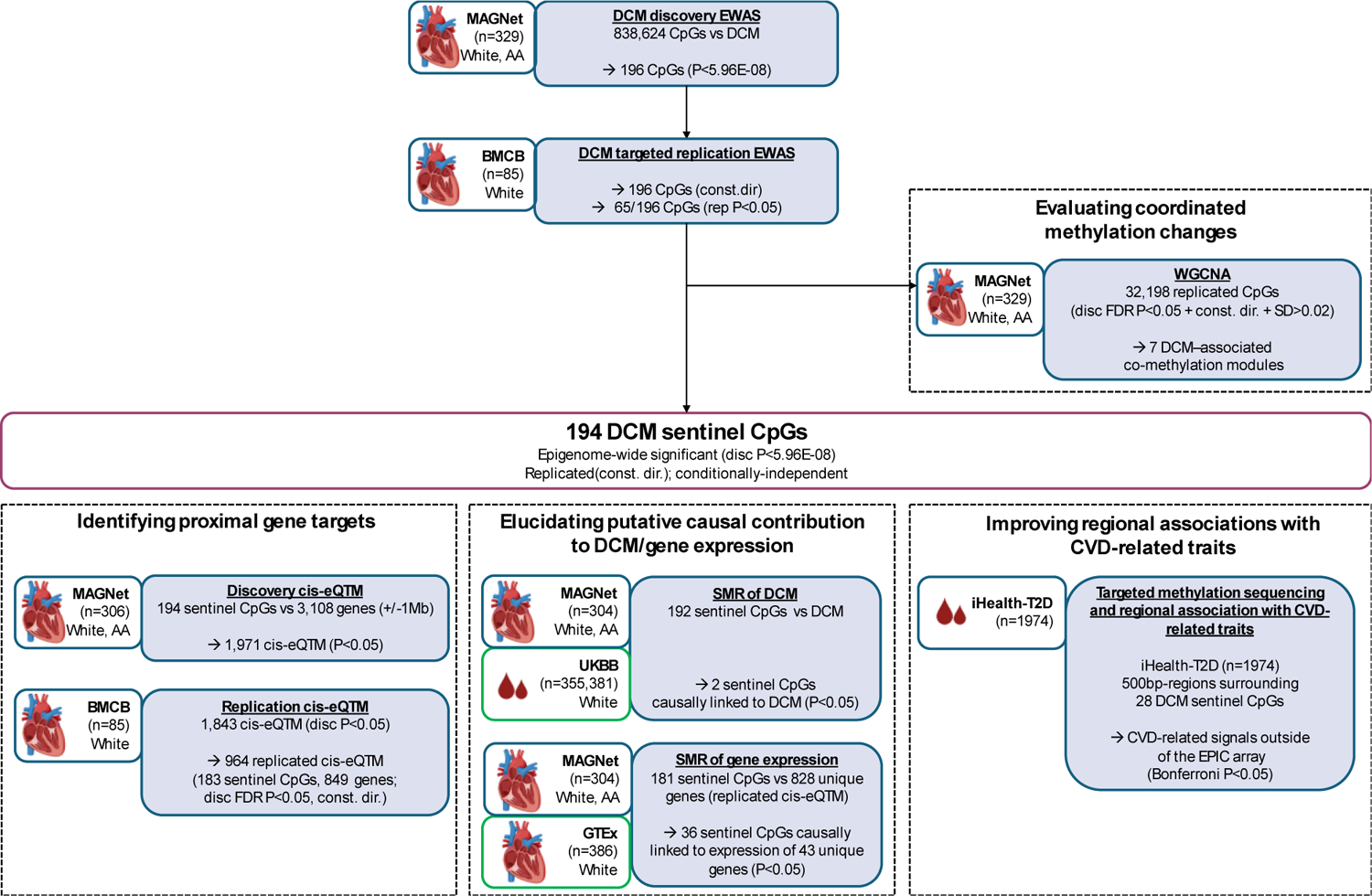
Study Design. Key abbreviations used: (Analyses) EWAS=epigenome-wide association study; eQTM=expression quantitative trait methylation analysis; (Ancestries) AA=African American; (Cohorts) MAGNet=Myocardial Applied Genomics Network; BMCB=Bruce McManus Cardiovascular Biobank; UKBB=UK Biobank.

### Epigenome-wide association analysis

We performed EWAS of DCM using genomic DNA extracted from LV free-wall tissue, with separate EWAS carried out for Whites and African Americans within the discovery cohort (MAGNet), followed by inverse-variance meta-analysis (Methods). From discovery-stage EWAS, 196 CpG sites were associated with DCM at a Bonferroni-corrected threshold of P<5.96E-08 (0.05/838,624) (Figure 2). Subsequent targeted replication testing in the BMCB cohort confirmed consistent directionality of effect size estimates for all discovery CpGs (Supplementary Table 2). The 196 CpG sites were distributed across 171 genetic loci, with 150/171(88%) genetic loci containing a single sentinel CpG and 21/171 (12%) loci containing two or more sentinel CpGs (Supplementary Figure 1, Supplementary Table 2). Conditional analyses at each locus identified a total of 194 robustly associated and conditionally independent signals (‘sentinel CpGs’), which were further analysed for functional relevance and causal contribution to DCM pathogenesis.

**Figure 2:**
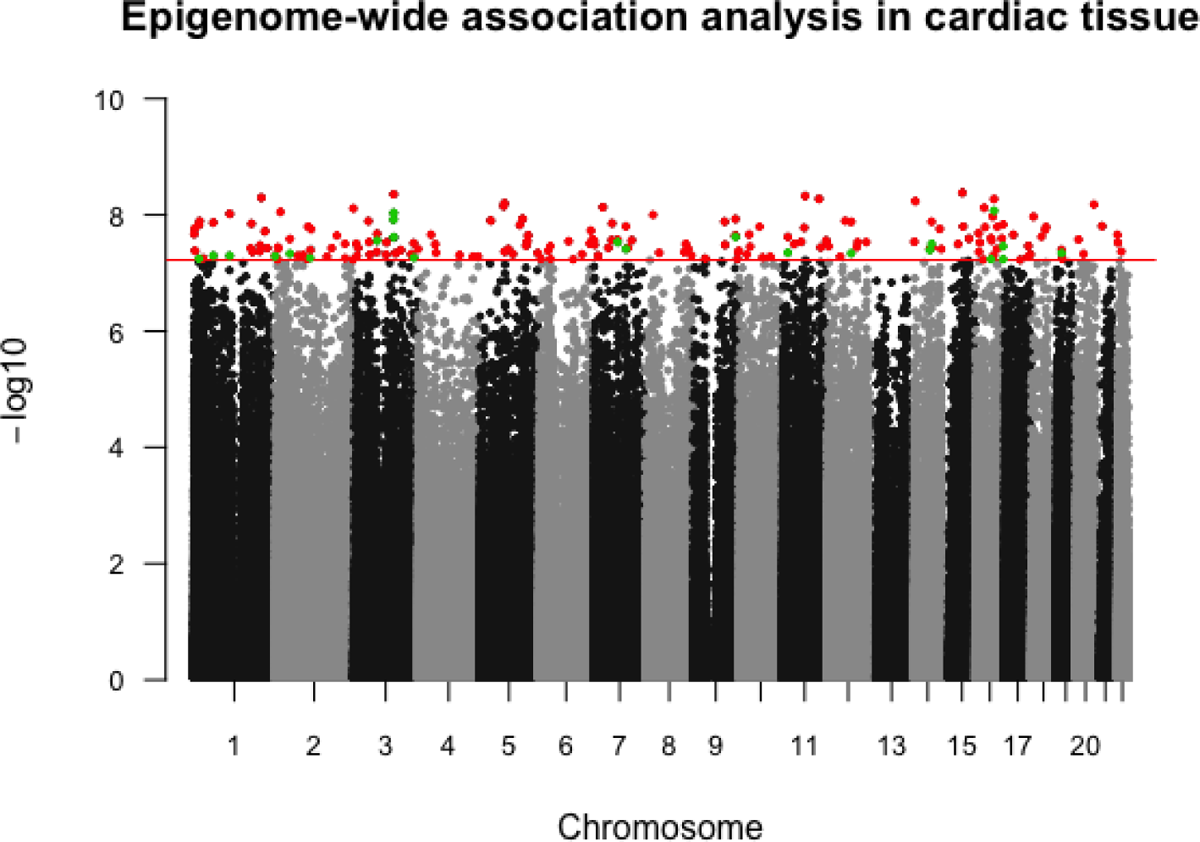
Manhattan plot for discovery-stage EWAS of DCM. The horizontal significance line (red) corresponds to the epigenome-wide significance threshold (P<5.96E-08, 0.05/838,624 tests). The 194 DCM sentinel CpGs are highlighted. At genomic loci with >1 epigenome-wide significant signal, secondary signals (green) were identified by conditioning on the lead signal (lowest P in region; red).

Unsupervised hierarchical clustering based on the methylation levels of the 194 sentinel CpGs resulted in two distinct clusters, segregating samples by their respective case and control status. The ‘case’ cluster correctly consisted of 128 (88%) cases, while the ‘control’ cluster had 153 (83%) controls (binomial P<2.2E-16) (Figure 3). This finding supports a perturbation of DNA methylation in DCM. Clustering by case and control status persisted in ancestry-specific unsupervised hierarchical clustering of methylation levels of the sentinel CpGs (Supplementary Figure 2).

**Figure 3:**
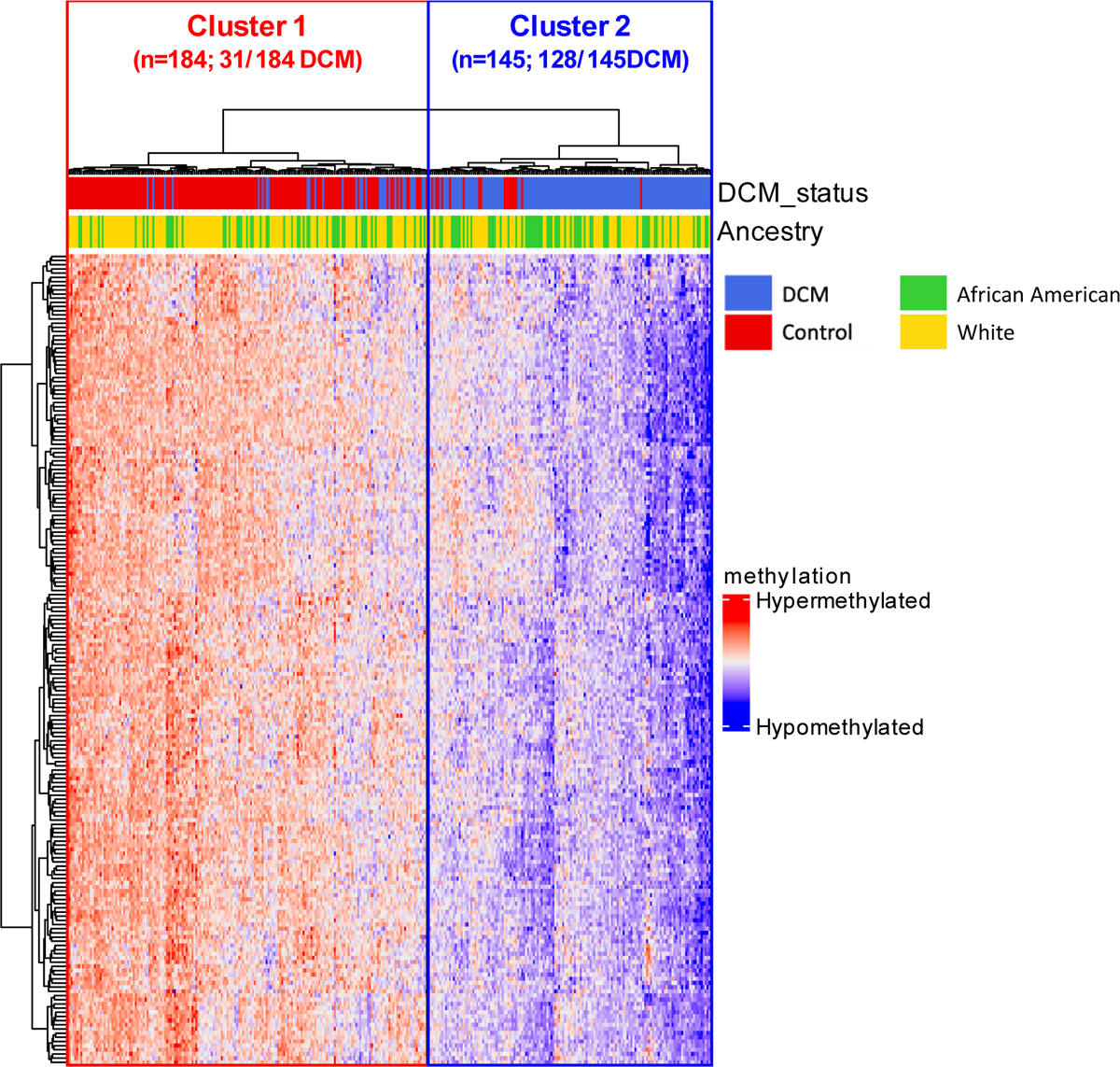
Unsupervised clustering using methylation levels of 194 sentinel CpGs. DCM, dilated cardiomyopathy.

### Enrichment of Sentinel CpGs in active gene regulatory regions and impact on proximal gene expression

To understand the regulatory role of sentinel CpGs, we first examined the enrichment of annotated chromatin states. Sentinel CpGs were enriched in transcriptionally active chromatin states and depleted in repressed Polycomb states of LV tissue (permutation test *P* <0.001), compared to a background of CpGs matched by relationships to gene and CpG islands (Supplementary Figure 3A). Consistent with their enrichment in active LV chromatin states, sentinel CpGs were also enriched for location in H3K4me3-marked regions, typically representing transcriptionally active chromatin and promoters (permutation test P <0.001) (Supplementary Figure 3B).

To identify sentinel CpGs that impacted proximal gene expression, hereon referred to as expression quantitative methylation loci (eQTM), we examined the association between methylation levels of the 194 sentinel CpGs and expression of their proximal genes (<1Mb) present in the discovery (MAGNet) and replication (BMCB) RNAseq datasets. We found the 194 sentinel CpGs to be enriched for association with proximal gene expression in LV tissue (3.80-fold compared to expectations under the null hypothesis; *P* <0.001) (Supplementary Figure 4). Subsequent targeted replication testing on sentinel CpG-gene pairs reaching FDR *P* <0.05 in discovery-stage association testing confirmed consistent directionality of effect size estimates between 183 sentinel CpGs and 849 unique proximal genes (‘replicated eQTMs’) (Supplementary Table 3).

### Summary data-based Mendelian Randomisation to infer disease causation

Having obtained initial evidence of sentinel CpG contribution to transcriptional regulation, we next investigated whether these CpGs had a causal relationship with both DCM and proximal gene expression using Summary data-based Mendelian Randomization (SMR). Separate causal analyses were conducted for DCM and gene expression. We found two sentinel CpGs that were causally linked to DCM (cg08140459 and cg12359658; p<0.05), with subsequent validation testing via one-sample MR confirming consistent directionality of causal estimate for cg08140459-DCM (p<0.05) (Supplementary Table 4).

To gain further insight into the molecular mechanisms underpinning the contribution of sentinel CpGs to DCM, we conducted a separate SMR of gene expression, focusing on replicated eQTMs. After excluding sentinel CpGs without suitable instrumental variables (e.g., meQTL not included in public (GTEx) dataset), 931 eQTMs (181 unique sentinel CpGs, 828 unique genes) could be analysed in the SMR of gene expression. Out of the 181 sentinel CpGs analysed, 36 sentinel CpGs showed putative causal relationships with 43 unique proximal genes (p<0.05) (Supplementary Table 5).

We further integrated evidence from multi-omics analyses to identify the most relevant sentinel CpGs for DCM pathogenesis. Among the 36 sentinel CpGs causally linked to proximal gene expression, we selected the three CpGs with the highest posterior probability for a shared causal variant influencing both CpG methylation and proximal gene expression (cg09862509-*IER5*, coloc.abf-PP.H4 = 0.91; cg11793257-*ENTPD6*, coloc.abf-PP.H4 = 0.69; cg11793257-*ABHD12*, coloc.abf-PP.H4 = 0.47; cg06807905*-KCNC4*, coloc.abf-PP.H4 = 0.51) (Figure 4, Supplementary Figure 5). Causal estimates for these CpGs were validated in a one-sample MR setting, showing consistent direction of association (Supplementary Data 5).

**Figure 4.**
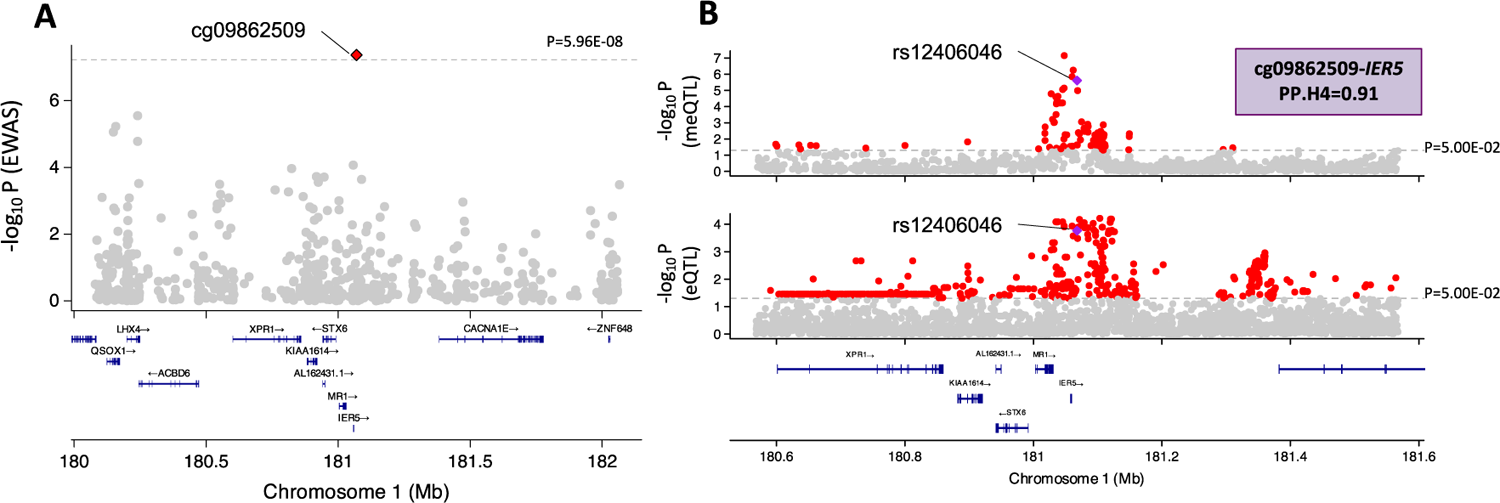
Regional plots for the sentinel CpG gene pair showing the highest posterior probability of colocalization. Two types of regional association plots are shown for cg09862509-*IER5*: **A. Associations of CpGs surrounding cg09862509 with DCM from discovery-stage EWAS (MAGNet).** The displayed region is centered on cg09862509 (red; bordered) B. Genetic associations for cg09862509 and *IER5*. Genetic associations were obtained for a +/-500kb region centered on the instrumental variable used in SMR assess causal relationships between cg09862509 methylation and *IER5* gene expression (meQTL from MAGNet; eQTL from GTEx.) Posterior probability of colocalization between cg09862509 meQTLs and *IER5* eQTLs was 0.91. Gene tracks for all regional plots consist of protein-coding genes obtained from Ensembl Release 75 (last release based on hg19 assembly).

Although the cardiac roles of the target genes have not been well-studied, existing investigations suggest functions with potential relevance to DCM pathogenesis. *IER5* is a transcription factor regulating cell proliferation, possibly via the crosstalk between Notch and DNA damage response pathways. *ENTPD6* is a nucleotide-metabolising enzyme that is predominantly expressed in the heart and functions in platelet recruitment and aggregation.^17^ The potassium ion channel *KCNC4* influences neural cell survival and apoptosis under oxidative stress conditions in mice and may also play a role in regulating myocardial action potential.^18,19^ *ABDH12* is an enzyme that hydrolyses endocannabinoids, a class of lipids regulating a wide range of pathologies including platelet aggregation, vasodilation, and the maintenance of energy balance.^20^

### Genes mapped to CpG sites demonstrating coordinated changes in methylation patterns are enriched in disease-relevant pathways

Beyond analysing single CpG associations, examining coordinated methylation changes across multiple CpG sites and their linked genes could reveal disease-relevant pathways regulated by DCM methylation. To achieve this, we conducted weighted correlation network analysis (WGCNA), constructing co-methylation modules using methylation levels of 32,918 DCM-associated CpG sites (discover EWAS FDR *P*<0.05, with consistent direction of association in replication EWAS, SD> 0.02 across all samples). Seven co-methylation modules were identified (Figure 5). Regressing the module eigengene (the first principal component of module-specific methylation intensities) against DCM confirmed an association with DCM for all modules (Supplementary Table 6).

**Figure 5.**
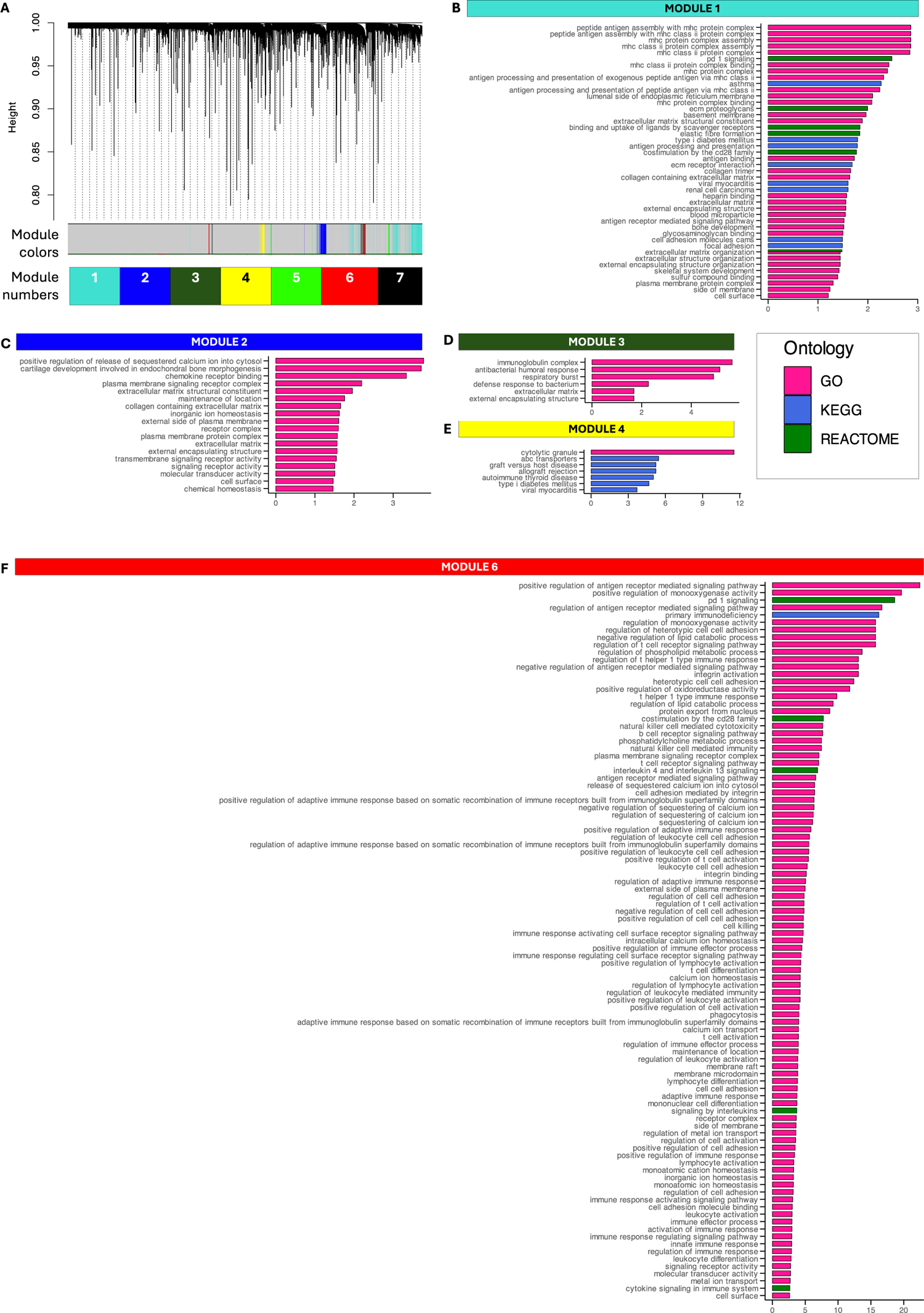
Co-methylation modules identified using DCM-associated CpGs. A. Hierarchical cluster tree (dendogram) of co-methylation modules. WGCNA identified seven modules that exhibit conservation between co-methylation networks constructed independently within the White (n=209) and African American (n=118) ancestries. The color band underneath the tree indicates distinct modules (grey =CpG sites that are not clustered into any module). **B-F. Bar plots of over-represented gene sets (FDR P<0.05) in co-methylation modules.** Genes were assigned to CpGs that were identified as replicated cis-eQTMs, signifying a strong correlation between methylation and gene expression (MAGNet FDR P<0.05, consistent direction of association in BMCB analysis). Gene set enrichment within a module was assessed against a background set consisting of all genes mapped to CpG sites that were used to identify co-methylation patterns (9,321 unique genes). Enriched gene sets are displayed by decreasing order of fold-change (x-axis; calculated as the ratio of the proportion of overlapping genes linked to module-specific CpGs to the proportion of overlapping genes linked to background CpGs) GO = gene ontology; KEGG = Kyoto Encyclopedia of Genes and Genomes.

To investigate the biological relevance of co-methylation modules, we looked for over-represented gene sets and enrichment in binding sites of known transcription factors (TFs). (Figure 5, Supplementary Figure 6) Gene set overrepresentation was analyzed using genes belonging to replicated eQTM pairs of module-specific CpGs.

Multiple modules were enriched in gene sets associated with cardiac development and key pathological processes in heart failure, such as inflammation and alterations in extracellular matrix (ECM) structure and composition (Figure 5, Supplementary Table 7). Notably, Module 5 CpG loci were enriched in binding sites of *YAP*, a key effector of the Hippo pathway that has a well-established and essential role in cardiac development^21^(Supplementary Figure 6). Certain modules showed enrichment for gene sets and transcription regulators more specific to cardiomyopathy. For instance, Module 1 was enriched for gene sets related to muscle adaptation, including processes such as hypertrophy or remodeling in response to injury (Figure 5). Correspondingly, Module 1 CpG loci were enriched in binding sites of TFs regulating cardiomyocyte hypertrophy, such as *THRA, PTRF, NSD2, HOXA9,* and *LZBTB* (*P*<0.001) (Supplementary Figure 6). Module 6 was enriched for gene sets related to nucleotide and lipid metabolism, regulation of calcium ion transport and homeostasis, as well as enzymatic activity catalyzing chemical modification of nucleic acids and other biomolecules (Figure 5). Supporting the enrichment of metabolism-related gene sets, binding sites of *RXR*, a transcription factor known to regulate cardiac metabolism, was enriched amongst Module 6 CpG loci (P<0.001)^23^ (Supplementary Figure 6).

### Fine-mapping sentinel CpGs to investigate regional associations with traits related to cardiac disease and disease risk

As the methylation array covers only 2-3% of CpG sites in the epigenome, we sought to improve our investigation of regional associations using an existing target methylation sequencing dataset to increase coverage of CpG sites (+/- 500bp) surrounding the top-performing DCM sentinel CpGs. Targeted methylation sequencing was performed using blood samples of individuals from the iHealth-T2D study, which is well phenotyped for various traits relevant to CVD (n=1974). We assessed regional associations with (i) CVD such as myocardial infarction, angina, and coronary heart disease; (ii) indicators of CVD risk including blood pressure (systolic and diastolic), Framingham Coronary Heart Disease score, and hypertension/history of hypertension; (iii) high-sensitivity C-reactive protein (hsCRP), an inflammatory marker and predictor of severe cardiac outcomes in individuals without prior cardiac disease, and (iv) creatinine, a renal marker reflecting renal function that can worsen in heart failure.

Targeted sequencing was performed on regions surrounding 28 DCM sentinel CpGs (28 regions) (Supplementary Table 8). A total of 293 CpG sites were captured, with two to 24 CpG sites captured in each region. Seventeen regions had significant associations with at least one of the 10 unique CVD traits (Bonferroni-corrected P<0.05) (Supplementary Table 9). At 14 regions, the CpGs with the strongest association with our CVD traits were not CpGs on the EPIC array, further illustrating the added value brought upon by targeted sequencing. Multiple independent signals were also found for two regions, whereby conditioning on the lead signal revealed secondary signals in both regions.

To illustrate the utility of targeted sequencing to improve regional associations with DCM, we highlight regions surrounding two DCM sentinel CpGs that contained signals for CVD-relevant traits (Bonferroni-corrected P<0.05) from CpGs not present on the EPIC array (Figure 6A-C). In the region surrounding cg13173392 (EWAS P=4.71E-09; third most significant hit), targeted methylation sequencing identified stronger CHD and DBP signals from CpGs that were correlated with cg13173392 (chr11_69245749 with CHD, *P* = 1.93E-03, |r| = 0.46; chr11_69245240 with DBP, P=1.29E-03, |r| = 0.24) (Figure 6A,B), while cg13173392 itself was only nominally significant for both CHD (P=2.26E-01) and DBP (P=1.53E-01).

**Figure 6.**
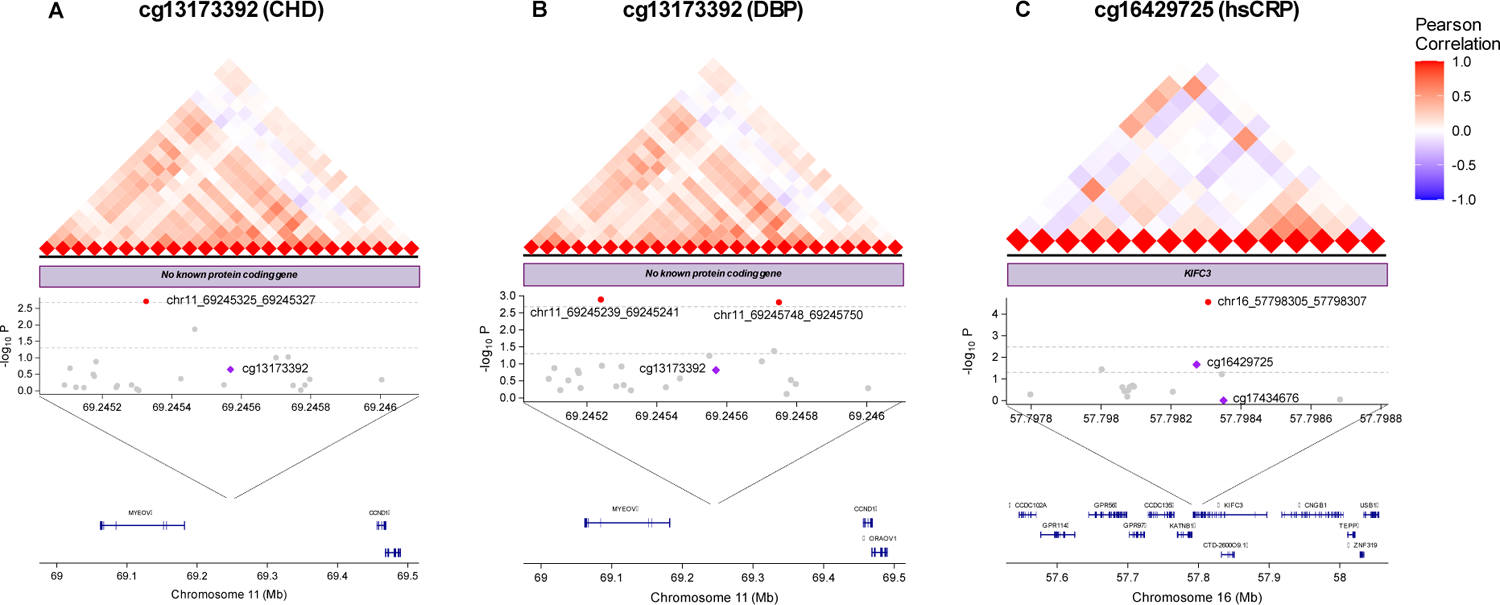
Regional associations of top DCM sentinel CpGs with CVD-related traits. Regions surrounding DCM sentinel CpGs (+/-500bp) were fine-mapped to discover signals for CVD-related trait using blood samples from a population-based cohort (iHealth). To highlight the value of fine-mapping disease-associated loci, these plots illustrate the regional associations for cg13173392 and cg16429725, which are the sentinel CpGs with the strongest associations with DCM amongst those that underwent fine-mapping. Three types of plots are included in each panel: (top) pairwise correlation in methylation levels between CpGs within a region; (middle) regional associations between CVD traits and CpGs; (bottom) protein-coding genes in the region. **A. Regional associations with coronary heart disease at the cg13173392 locus.** This region consists of 24 CpGs. The lead signal is chr11_69245325_69245327 (P=1.93 E-03). **B. Regional associations with diastolic blood pressure at the cg13173392 locus.** This region consists of 24 CpGs. The lead signal is chr11_69245239_69245241 (P=1.29 E-03). A secondary signal chr11_69245748_69245750 (P=1.55E-03) was revealed after conditioning on the lead signal. **C. Regional associations with the inflammatory mark high-sensitivity C-reactive protein at the cg16429725 locus.** This region consists of 15 CpGs. The lead signal is chr16_57798305_57798307 (P=2.79E-05). purple = DCM sentinel CpGs; red = neighbouring CpGs associated with the respective CVD traits (Bonferroni P<0.05).

In a separate region surrounding cg16429725 (EWAS P=5.33 E-09, sixth most significant hit), a CpG correlated with cg16429725 was identified as a strong hsCRP signal (chr16_57798306, P=2.79E-05, |r| = 0.57; Figure 6C), while cg16429725 itself was only nominally significant for hsCRP (P=2.16E-02). The investigated region surrounding cg16429725 is within the *KIFC3* gene involved in the formation, maintenance, and remodeling of the bipolar mitotic spindle. Although *KIFC3* has not been studied in a cardiac context, its potential contribution to inflammation is supported by its contribution to the hepatocellular carcinoma progression via the PI3K/AKT pathway and its role in stabilizing E-cadherin.^24,25^

We next combined methylation information from multiple CpGs in each region using a weighted methylation risk score to investigate associations with CVD. Of the 28 sequenced regions, 25 regions had significant methylation risk scores (Bonferroni P<0.05) for at least one investigated CVD trait. In 23 of these 25 regions, individual CpGs were not significantly associated with CVD (Bonferroni P<0.05). However, combining the methylation patterns of multiple CpGs within these regions into an MRS revealed significant associations with at least one of the investigated CVD traits, indicating that the combined effect of multiple CpGs provides stronger association with CVD traits than individual CpGs alone (Supplementary Table 10).

## DISCUSSION

We perform the largest EWAS of DCM in cardiac tissues to date (discovery n = 159 DCM, 170 control), extending on previous EWAS of DCM in terms of sample size and coverage of CpG sites. We identified and replicated 194 independent CpG signals for DCM that reached epigenome-wide significance (P<5.96E-08), with comprehensive multi-omics and causal analyses suggesting the causal contribution of a subset of our sentinels to DCM pathogenesis and transcriptional regulation. Fine-mapping of putative methylation markers and network analysis of coordinated changes across multiple DCM-associated CpG loci supported the relevance of DCM-linked methylation changes to cardiac development, disease pathogenesis as well as early indicators of cardiac risk.

Before the current investigation, the largest existing EWAS of DCM was conducted in a predominantly White cohort (discovery n=41 DCM, 31 control) using the older 450k array. This study used a staged multi-omics approach to identify CpG sites with conserved DCM-linked methylation alterations and impact on proximal gene expression.^9^ Among these, cg24884140 stood out as a promising diagnostic biomarker, showing consistent dysmethylation in both heart and blood, a strong link to gene expression, and outperforming the clinical standard heart failure biomarker N-terminal pro-BNP in ROC analysis for detecting DCM. Although cg24884140 did not qualify as one of the 194 sentinel CpGs in the current investigation, it was associated with DCM in the discovery-stage EWAS (beta=-12.67, P=8.70E-04), with the directionality of the association confirmed in subsequent replication testing (beta=-53.10, P=1.19E-02).

With individual-level genotype and transcriptome data available in addition to DNA methylation data, we could conduct multi-omics analyses that supported causal involvement of certain sentinel CpGs in DCM pathogenesis. The 194 sentinel CpGs were enriched in active chromatin states in heart tissue and significantly correlated with proximal gene expression more than expected by chance, supporting a role in transcriptional regulation. These preliminary indications of disease relevance probed us to investigate the putative causal contribution of sentinel CpGs to DCM. While individual-level genotype data enables the application of one-sample Mendelian Randomization to address population heterogeneity, Summary data-based Mendelian Randomisation (SMR) leverages data from large-scale association studies with higher statistical power to derive causal estimates. Therefore, we opted to conduct SMR as the primary analysis and one-sample MR to validate SMR causal estimates reaching nominal significance (P<0.05). We show putative causal relationships between cg08140459 and DCM. In our study, cg08140459 was robustly linked to the expression of *LTBP2*, a recently discovered prognostic biomarker for DCM, in independent cohorts.^26^ To gain insight into the molecular pathways in DCM pathogenesis that involve sentinel CpGs, we conducted a separate SMR analysis for gene expression, revealing the putative contribution of three sentinels to nearby genes (cg09862509-*IER5*, cg11793257-*ENTPD6*, cg11793257-*ABHD12*, cg06807905-*KCNC4*) with functions relevant to DCM pathogenesis, thus warranting further investigation in a cardiac context.

Using targeted methylation sequencing data from a population-based cohort phenotyped for multiple cardiac traits, we discovered independent CVD risk factor signals near DCM sentinels that were not captured by the EPIC array. While individual CpGs might demonstrate modest associations with CVD traits, combining methylation data from CpGs on a regional level strengthened these associations. This illustrates the value of targeted methylation sequencing in uncovering loci relevant to CVD traits.

Expanding our analysis beyond individual CpG associations, we employed co-methylation network analysis using CpGs associated with DCM in our discovery cohort (FDR P <0.05) to uncover pathways affected by methylation alterations in DCM. While gene sets associated with general cardiac pathologies such as inflammation and ECM alterations were enriched across multiple modules, gene sets related to muscle adaptation, metabolism, and calcium ion transport, which are more closely linked to cardiomyopathy were only enriched in specific modules. These findings indicate that CpG loci associated with DCM in our study regulate broad cardiac pathologies as well as those specific to cardiomyopathy Our study has some limitations. Firstly, the cell type heterogeneity of left ventricular tissue makes it challenging to delineate the specific cell types driving the associations between CpG methylation and DCM. Although bioinformatics cell type deconvolution methods exist for this task, the lack of reference methylation profiles for heart cell types means that only reference-free approaches can be applied. While reference-free deconvolution algorithms can predict distinct cell classes using major variations in methylation profiles, finding a biological basis to justify assigning these output classes to specific heart cell types (e.g. cardiomyocytes or cardiofibroblasts) currently poses a significant challenge. As single-cell profiling techniques for methylation and gene expression in cardiac cell types advance, future methylation studies of DCM should prioritise elucidating the cell type specificity of DCM-linked CpG methylation and the genes they regulate. Concerning our MR analyses, one limitation would be the potential bias in the causal relationships estimated by two-sample MR owing to population heterogeneity between the multi-ancestry MAGNet cohort (White, African American) used to generate meQTL and the predominantly White cohorts in which the GWAS of DCM and left ventricular eQTL analyses were conducted. Despite this, we did not restrict the meQTL analysis to only samples of White ancestry in our cohort with methylation and genotype data available (n=194) to maximise power for detecting left ventricular meQTL associations. On a related note, a second limitation of the MR analyses would be the limited sample size of the cohort used to generate meQTL, despite the current meQTL analysis being the largest to be conducted in left ventricular tissue to date.

Nonetheless, our study has important strengths. Besides being the largest existing study of DCM-linked methylation disturbances in left ventricular tissue to date, the current investigation extends previous methylation studies of DCM by seeking pevidence for the causal contribution of DCM-linked sentinel CpGs and by investigating regional associations with traits indicative of cardiac risk.

## CONCLUSION

This is the largest investigation of perturbed CpG methylation in DCM to be conducted in disease-relevant left ventricular tissue obtained from patients and controls. We identify CpGs independently and robustly associated with DCM and suggest molecular players in new, putative causal mechanisms by which DNA methylation may impact DCM. We also provide preliminary indication of the prognostic potential of DCM-linked CpGs associated with cardiac-relevant traits preceding overt disease in healthy individuals.

## Data Availability

Correspondence and requests for materials should be addressed to Marie Loh

## ACKNOWLEDGEMENTS AND FUNDING

This research is supported by a Start-Up Grant (awarded to Marie Loh [PI]) from Lee Kong Chian School of Medicine, Nanyang Technological University, Singapore.

